# Sociodemographic determinants of maternal health indicators in conflict-affected counties of Kenya: secondary analysis of data from the 2022 Kenya demographic and health survey

**DOI:** 10.64898/2026.04.22.26351520

**Authors:** Brigitte N Wandji, Charles O. Olungah, Eunice Atsali, Anne-Beatrice Kihara, Kireki Omanwa, Moses M Obimbo, Julius Ogeng’o

## Abstract

**Objective:** To analyse sociodemographic determinants of maternal health indicators in Kenya’s conflict-affected regions.

**Methods:** A cross-sectional secondary analysis of the 2022 Kenya Demographic and Health Survey (KDHS) was conducted. Conflict-affected counties were identified using ACLED (>25 fatalities). The sample included 1,060 women aged 15–49 years. Outcomes were adequate antenatal care (ANC 4+), facility delivery, and skilled birth attendance (SBA). Predictors included age, education, wealth, employment, residence, and county; intimate partner violence was adjusted for. Weighted descriptive statistics, chi-square tests, and multivariable logistic regression were applied (p<0.05).

**Results:** Six counties met conflict criteria. While 90.2% of women attended at least one ANC visit, only 53.5% achieved ANC 4+. Facility delivery and SBA were 68.2% and 72.2%, respectively. Adolescents (15–19) were least likely to attain adequate ANC; women aged 20–24 had higher odds (aOR=1.83; 95% CI: 1.01–3.34). Education strongly predicted outcomes: higher education increased ANC 4+ (aOR=2.74; 95% CI: 1.19–6.34) and facility delivery (aOR=2.72; 95% CI: 1.15–6.47). Wealth showed strong gradients: middle quintile increased facility delivery (aOR=5.50; 95% CI: 2.14–14.14), while richer quintile increased SBA (aOR=11.04; 95% CI: 2.06–59.25). Rural residence reduced facility delivery (aOR=0.32) and SBA (aOR=0.22). County disparities persisted. IPV was not independently associated.

**Conclusion:** Maternal health indicators in conflict-affected Kenya follow a marked inequity gradient. Adolescents, rural residents, and socioeconomically disadvantaged women are most excluded. Strengthening adolescent ANC continuity, reducing rural access barriers, and investing in education and economic empowerment are critical for improving outcomes.

## 1. Introduction

Maternal health outcomes remain among the most sensitive indicators of population health, health systems performance and social equity. Improvements in these indicators typically reflect a reduction in maternal morbidity and mortality. Yet, across many low- and middle-income settings, the benefits of maternal health progress are not experienced uniformly(1–3). Systemic and avoidable inequities that are shaped by the sociodemographic and economic realities of the women exist widely. These inequities undermine women’s ability to access and use essential services during pregnancy and childbirth and ultimately determine who receives timely, adequate care and who does not (3–5).

Maternal health indicators like adequate antenatal care (ANC) attendance, facility-based delivery, and skilled birth attendance (SBA) are critical indicators because they reflect continuity across the maternal care pathway (6). Their inadequacy or absence represent missed opportunities for early risk identification, timely referral, and skilled intrapartum care; thereby increasing the likelihood of severe complications. Structural inequities in access and use of maternal health services will be reflected in these indicators which serve as established proxies for maternal survival and preventable death(5,7).

Conflict and insecurity intensify these inequities in ways that extend beyond routine health-system limitations. In conflict-affected settings, health infrastructure may be damaged or under-resourced, governance and service delivery weakened, skilled providers displaced, and routine referral and transport network systems broken(8,9). Insecurity also constrains mobility and increases the financial and social costs of seeking care, effectively turning distance into a larger barrier. During conflict, a woman’s personal and socioeconomic resources such as education and ability to pay for safer transport become more decisive in determining whether she can access life-saving services(5,9,10).

Fundamentally, sociodemographic factors shape maternal health indicators by determining women’s capacity to access, sustain, and benefit from essential services across the continuum of care (3,5). This influence is amplified in conflict-affected settings, where insecurity and system disruption magnify pre-existing inequalities (11). In addition, these patterns are not uniform; the interaction between conflict intensity, infrastructure stability, and local socioeconomic conditions produces variation across settings (12). Understanding these shifts in sociodemographic gradients within fragile environments is critical for informing targeted, sub-national planning and equity-driven maternal health interventions.

There’s however insufficient disaggregated evidence on how specific sociodemographic determinants influence maternal health indicators within conflict-affected counties in Kenya (13). This study therefore sought to analyse the sociodemographic factors that shape adequate antenatal care, facility delivery, and skilled birth attendance in Kenya’s conflict-affected regions.

## 2.0 Methodology

### 2.1 Study Design and Data Source

This study employed a descriptive, cross-sectional design utilizing secondary data from the 2022 Kenya Demographic and Health Survey (KDHS). The KDHS is a nationally representative survey implemented by the Kenya National Bureau of Statistics (KNBS) in collaboration with the Ministry of Health, with technical support from ICF through the USAID-funded DHS Program (14). The survey utilizes a two-stage stratified sampling design to provide estimates for key maternal health indicators at national and sub-national (county) levels.

### 2.2 Identification of Conflict-Affected Counties

Kenya has 47 counties (15). Of these 47, conflict-affected counties were identified using the Armed Conflict Location & Event Data (ACLED) database, an independent repository of disaggregated conflict events (16). Counties were classified as "conflict-affected" if they recorded significant organized violence resulting in more than 25 fatalities within the 24-month period preceding the KDHS data collection (17)

### 2.3 Study Population and Sampling

The study population consisted of women of reproductive age (15–49 years) residing in the conflict affected counties. For maternal care indicators (Antenatal Care, Facility Delivery and Skilled Birth Attendance), the analysis focused on 1,060 women who had a live birth in the two years preceding the survey.

### 2.4 Measurement of Variables

For Dependent Variables (Maternal Health Indicators), three primary outcomes were analysed; Adequate Antenatal Care (attendance of four or more ANC visits during the most recent pregnancy), Facility Delivery (Giving birth in a public, private, or faith-based health facility) and Skilled Birth Attendance (Delivery assisted by a doctor, nurse, or midwife).

The primary explanatory variables were sociodemographic characteristics derived from the KDHS dataset, including: maternal age, educational attainment (none, primary, secondary, higher), household wealth index (coded into quintiles: poorest, poorer, middle, richer, richest), employment status, place of residence (urban/rural) and county of residence.

Experiences of Intimate Partner Violence (IPV) were included as a covariate in the multivariable models to adjust for its potential confounding effect.

### 2.5 Statistical Analysis

Data analysis was performed using Stata. For descriptive statistics, frequencies and percentages were used to describe the sociodemographic profile of the sample. Chi-square tests were utilized to examine the associations between sociodemographic factors and maternal health indicators in bivariable analysis. Logistic regression models were fitted to estimate the adjusted odds ratios with 95% confidence intervals in multivariable analysis. Statistical significance was set at p<0.05.

### 2.6 Ethical Considerations

The study is a secondary analysis of de-identified KDHS data. Access to the dataset was granted by MEASURE DHS. No further ethical approval was required as the study involved no identifiable personal information, no direct human subject contact and used public-domain data.

## 3. Results

### 3.1 Conflict affected Counties

Six (6) counties out of 47 were identified from the database of the Armed Conflict Localization and Events Data (ACLED) as having significant conflict during the twenty-four months which spanned the period of interest over which the variables were being studied in the KDHS. The main conflict affected counties were Lamu, Mandera, Marsabit, Isiolo, Samburu and Elgeyo Marakwet.

### 3.2 Background (sociodemographic) characteristics of respondents

1060 respondents from the 6 conflict affected counties were included in the analysis for maternal care indicators (ANC 4+ visits, Health Facility delivery, and Skilled Birth Attendant-SBA.

The study population reflected marked social and geographic diversity across education level, household wealth, employment status, residence, and county of origin, underscoring the heterogeneous structural environments within conflict-affected settings.

Table 1 presents the background characteristics of respondents included in the analysis. A comparative majority were in their late 20s in terms of age and had no formal education. Majority of them were also in the poorest wealth quintile, resided in rural areas, lived in male-headed households and were not working.

**Table 1:** Background information of participants.

|  | ANC attendance, ANC 4+ visits, Health Facility delivery, and SBA (N=1060) | Met and Unmet needs (N=521) |
| --- | --- | --- |
| Variable | n (%) | n (%) |
| Age |  |  |
| 15-19 | 86(8.1) | 49(9.4) |
| 20-24 | 238(22.5) | 92(17.6) |
| 25-29 | 284(26.9) | 117(22.5) |
| 30-34 | 221(20.9) | 108(20.8) |
| 35-39 | 169(15.9) | 87(16.6) |
| 40-44 | 53(4.9) | 44(8.5) |
| 45-49 | 8(0.8) | 24(4.5) |
| **Educational level** |  |  |
| None | 515(48.6) | 223(42.8) |
| Primary | 265(25.0) | 136(26.0) |
| Secondary | 211(19.9) | 108(20.7) |
| Higher | 69(6.5) | 54(10.4) |
| **Wealth index** |  |  |
| Poorest | 609(57.5) | 268(51.5) |
| Poorer | 115(10.9) | 42(8.0) |
| Middle | 159(14.9) | 90(17.3) |
| Richer | 122(11.5) | 78(14.9) |
| Richest | 55(5.2) | 43(8.2) |
| **Residence** |  |  |
| Urban | 282(26.6) | 130(24.9) |
| Rural | 778(73.4) | 391(75.1) |
| **Sex of household head** |  |  |
| Male | 719(67.9) | 347(66.5) |
| Female | 341(32.1) | 174(33.5) |
| **Working status** |  |  |
| Not working | 845(79.7) | 384(73.8) |
| Working | 215(20.3) | 137(26.2) |
| **County** |  |  |
| Lamu | 85(8.1) | 46(8.8) |
| Mandera | 276(26.0) | 103(19.8) |
| Marsabit | 172(16.2) | 82(15.7) |
| Isiolo | 113(10.7) | 56(10.8) |
| Samburu | 197(18.6) | 92(17.6) |
| Elgeyo-Marakwet | 217(20.5) | 142(27.3) |

Nearly half of the respondents (48.6%) had no formal education, and a vast majority (57.5%) were in the "Poorest" wealth quintile. The population was predominantly rural (73.4%) and engaged in unpaid work or unemployed (79.7%). Mandera and Elgeyo Marakwet contributed the largest proportion of respondents (26.0% and 20.5%, respectively).

### 3.3 Determinants of Antenatal Care (ANC) Utilization

While the overall prevalence of attending at least one ANC visit was high (90.2%), the prevalence of adequate ANC (4+ visits) was significantly lower at 53.5%.

**Table 2:** Bivariable and multivariable analysis of factors associated with the odds 4+ ANC visits.

| Variables | p(95% CI)<br>4+ visits | $\chi^2$ (p-value) | cOR (95% CI), p-value | aOR (95% CI), p-value |
| --- | --- | --- | --- | --- |
| **Age** |  | 16.06(0.099) |  |  |
| 15-19 | 46.9(34.9-59.4) |  | Ref. | Ref. |
| 20-24 | 61.8(52.7-70.1) |  | **1.83(1.03-3.23),0.038** | **1.83(1.01-3.34),0.047** |
| 25-29 | 46.4(38.8-54.2) |  | 0.98(0.55-1.74),0.937 | 1.08(0.58-1.99),0.808 |
| 30-34 | 52.8(44.6-60.8) |  | 1.26(0.73-2.19),0.403 | 1.53(0.84-2.79),0.168 |
| 35-39 | 56.3(47.6-64.6) |  | 1.46(0.80-2.64),0.214 | 1.74(0.91-3.33),0.093 |
| 40-44 | 56.0(38.0-72.5) |  | 1.44(0.62-3.35),0.400 | 1.69(0.69-4.18),0.250 |
| 45-49 | 74.1(38.9-92.8) |  | 3.24(0.63-16.61),0.158 | 4.15(0.88-19.51),0.071 |
| **Educational level** |  | **19.23(0.012)** |  |  |
| None | 47.2(40.7-53.8) |  | Ref. | Ref. |
| Primary | 56.7(49.2-64.0) |  | 1.46(0.99-2.16),0.054 | 1.38(0.87-2.17),0.168 |
| Secondary | 59.6(49.9-68.7) |  | **1.64(1.03-2.65),0.039** | 1.61(0.95-2.75),0.078 |
| Higher | 69.2(55.4-80.2) |  | **2.51(1.32-4.77),0.005** | **2.74(1.19-6.34),0.019** |
| **Working status** |  | **8.29(0.011)** |  |  |
| Not working | 51.3(46.3-56.2) |  | Ref. | Ref. |
| Working | 62.2(55.1-68.9) |  | **1.57(1.11-2.21),0.011** | 1.21(0.79-1.83),0.373 |
| **Wealth index** |  | 9.03(0.152) |  |  |
| Poorest | 49.6(43.5-55.7) |  | Ref. |  |
| Poorer | 58.9(47.3-69.7) |  | 1.46(0.86-2.47),0.159 |  |
| Middle | 57.1(48.3-65.7) |  | 1.35(0.88-2.07),0.165 |  |
| Richer | 60.0(50.3-69.0) |  | 1.52(0.97-2.38),0.065 |  |
| Richest | 60.4(47.1-72.3) |  | 1.55(0.86-2.80),0.148 |  |
| **Sex of household head** |  | 1.44(0.402) |  |  |
| Male | 52.2(47.2-57.2) |  | Ref. |  |
| Female | 56.2(48.4-63.6) |  | 1.17(0.81-1.70),0.402 |  |
| **Residence** |  | 0.44(0.586) |  |  |
| Urban | 55.2(48.7-61.5) |  | Ref. |  |
| Rural | 52.9(47.6-58.1) |  | 0.91(0.65-1.27),0.586 |  |
| **Physical violence** |  | 0.47(0.523) |  |  |
| No | 54.0(49.3-58.6) |  | Ref. |  |
| Yes | 51.2(43.6-58.7) |  | 0.89(0.63-1.26),0.523 |  |
| **Emotional violence** |  | 1.24(0.319) |  |  |
| No | 52.6(47.9-57.3) |  | Ref. |  |
| Yes | 56.7(49.3-63.8) |  | 1.18(0.85-1.64),0.319 |  |
| **Sexual violence** |  | 0.83(0.464) |  |  |
| No | 53.3(48.9-57.5) |  | Ref. |  |
| Yes | 61.5(39.6-79.6) |  | 1.40(0.56-3.49),0.466 |  |
| **Overall IPV** |  | 0.94(0.399) |  |  |
| No | 52.6(47.5-57.5) |  | Ref. |  |
| Yes | 55.9(49.3-62.2) |  | 1.14(0.84-1.56),0.399 |  |
| **County** |  | **53.10(<0.001)** |  |  |
| Lamu | 68.8(60.8-75.7) |  | Ref. | Ref. |
| Mandera | 37.0(29.2-45.7) |  | **0.27(0.16-0.44),<0.001** | **0.39(0.21-0.73),0.003** |
| Marsabit | 67.4(56.3-76.8) |  | 0.94(0.52-1.69),0.832 | 1.28(0.67-2.44),0.461 |
| Isiolo | 50.6(41.5-59.7) |  | **0.47(0.28-0.77),0.003** | **0.49(0.29-0.82),0.007** |
| Samburu | 56.9(47.2-66.1) |  | 0.60(0.36-1.01),0.056 | 0.74(0.42-1.29),0.282 |
| Elgeyo-marakwet | 55.8(45.7-65.5) |  | **0.57(0.34-0.98),0.043** | **0.51(0.29-0.89),0.019** |
| cOR=crude odds ratio, aOR=adjusted odds ratio, Ref=Reference, CI=Confidence interval, |  |  |  |  |

### 3.4 Structural Barriers to Facility Delivery

The utilization of facility-based delivery (68.2%) showed the sharpest sociodemographic gradients, particularly regarding wealth and geographic residence.

**Table 3:** Bivariable and multivariable analysis of factors associated with the odds of facility delivery.

| Variables | PoD p(95% CI) | $\chi^2$ (p-value) | cOR (95% CI), p-value | aOR (95% CI), p-value |
| --- | --- | --- | --- | --- |
| **Age** |  | **18.57(0.039)** |  |  |
| 15-19 | 65.9(53.1-76.8) |  | Ref. |  |
| 20-24 | 76.2(68.3-83.0) |  | 1.65(0.87-3.13),0.124 |  |
| 25-29 | 72/5(66.5-77.8) |  | 1.36(0.77-2.42),0.289 |  |
| 30-34 | 61.7(53.8-69.1) |  | 0.83(0.44-1.59),0.576 |  |
| 35-39 | 62.2(51.7-71.6) |  | 0.85(0.46-1.57),0.599 |  |
| 40-44 | 60.7(41.4-77.2) |  | 0.79(0.30-2.21),0.645 |  |
| 45-49 | 57.4(22.8-86.0) |  | 0.69(0.14-3.58),0.663 |  |
| **Educational level** |  | **240.29(<0.001)** |  |  |
| None | 45.7(39.2-52.2) |  | Ref. | Ref. |
| Primary | 85.7(80.3-89.9) |  | **7.16(4.47-11.47),<0.001** | **2.09(1.09-3.99),0.026** |
| Secondary | 90.9(84.0-95.0) |  | **11.83(5.79-24.14),<0.001** | **2.72(1.15-6.47),0.023** |
| Higher | 100.0 |  | Omitted | omitted |
| **Working status** |  | **51.43(<0.001)** |  |  |
| Not working | 63.8(58.0-67/8) |  | Ref. | Ref. |
| Working | 88.5(80.0-93/7) |  | **4.53(2.28-9.04),<0.001** | 1.22(0.59-2.53),0.596 |
| **Wealth index** |  | **244.86(<0.001)** |  |  |
| Poorest | 49.0(43.1-54.9) |  | Ref. | Ref. |
| Poorer | 90.4(82.4-95.0) |  | **9.86(4.76-20.45),<0.001** | **3.55(1.50-8.37),0.004** |
| Middle | 94.9(88.9-97.7) |  | **19.21(8.21-44.98),<0.001** | **5.50(2.14-14.14),<0.001** |
| Richer | 95.3(86.5-98.5) |  | **21.07(6.53-67.95),<0.001** | **4.51(1.24-16.44),0.023** |
| Richest | 97.4(82.8-99.7) |  | **38.69(4.93-303.33),0.001** | 3.39(0.36-31.64),0.281 |
| **Sex of household head** |  | 0.68(0.458) |  |  |
| Male | 69.0(64.4-73.3) |  | Ref. |  |
| Female | 66.5(59.2-73.1) |  | 0.89(0.66=1.22),0.458 |  |
| **Residence** |  | **68.32(<0.001)** |  |  |
| Urban | 87.9(82.1-92.0) |  | Ref. | Ref. |
| Rural | 61.1(55.5-66.4) |  | **0.22(0.13-0.36),<0.001** | **0.32(0.15-0.67),0.003** |
| **Physical violence** |  | 0.17(0.694) |  |  |
| No | 67.9(63.1-72.4) |  | Ref. |  |
| Yes | 69.5(61.6-76.4) |  | 1.08(0.75-1.55),0.694 |  |
| **Emotional violence** |  | 0.92(0.450) |  |  |
| No | 67.5(62.5-72.1) |  | Ref. |  |
| Yes | 70.8(62.4-78.0) |  | 1.17(0.78-1.75),0.451 |  |
| **Sexual violence** |  | 0.04(0.851) |  |  |
| No | 68.1(63.7-72.3) |  | Ref. |  |
| Yes | 69.9(49.0-84.9) |  | 1.09(.46-2.58),0.851 |  |
| **Overall IPV** |  | 1.20(0.334) |  |  |
| No | 67.2(62.2-71.9) |  | Ref. |  |
| Yes | 70.7(63.8-76.9) |  | 1.18(0.84-1.64),0.334 |  |
| **County** |  | **184.99(<0.001)** |  |  |
| Lamu | 92.0(84.5-96.0) |  | Ref. | **Ref.** |
| Mandera | 51.0(41.5-60.4) |  | **0.09(0.04-0.21),<0.001** | **0.24(0.08-0.79),0.020** |
| Marsabit | 55.3(44.0-66.0) |  | **0.11(0.05-0.26),<0.001** | **0.32(0.11-0.94),0.038** |
| Isiolo | 87.4(77.8-93.3) |  | 0.61(0.22-1.67),0.331 | 0.98(0.27-3.37),0.978 |
| Samburu | 52.9(40.6-64.9) |  | **0.09(0.04-0.24),<0.001** | **0.28(0.09-0.78),0.016** |
| Elgeyo-Marakwet | 94.9(89.0-97.7) |  | 1.61(0.53-4.88),0.400 | 1.76(0.53-6.04),0.346 |
| **cOR=crude odds ratio, aOR=adjusted odds ratio, Ref=Reference, CI=Confidence interval,** |  |  |  |  |

### 3.5 Structural Barriers to Skilled Birth Attendance (SBA)

The utilization of skilled birth attendance (72.2%) showed the sharp sociodemographic gradients. Patterns for skilled birth attendance mirrored those observed for facility delivery.

**Table 4:**
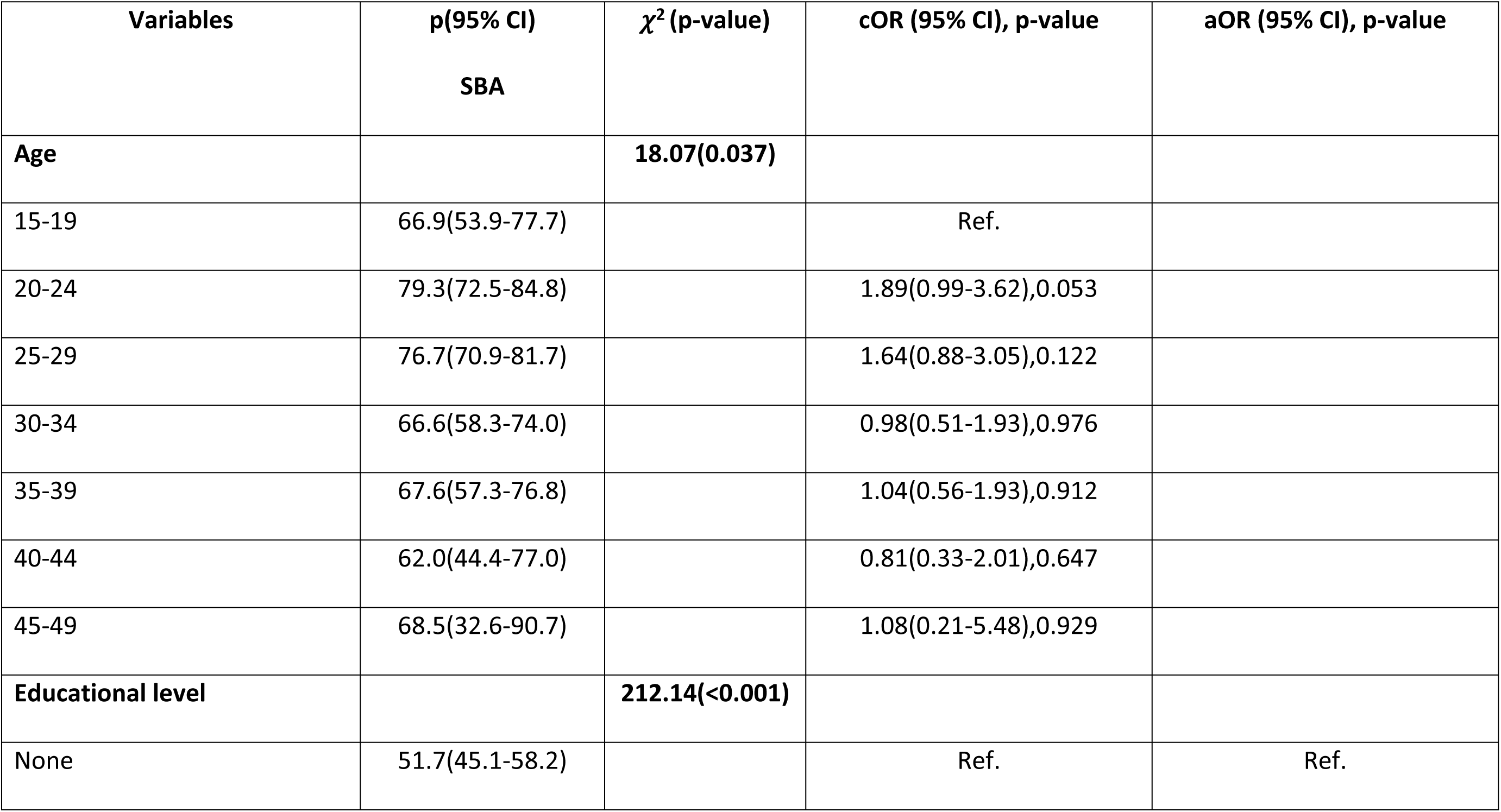
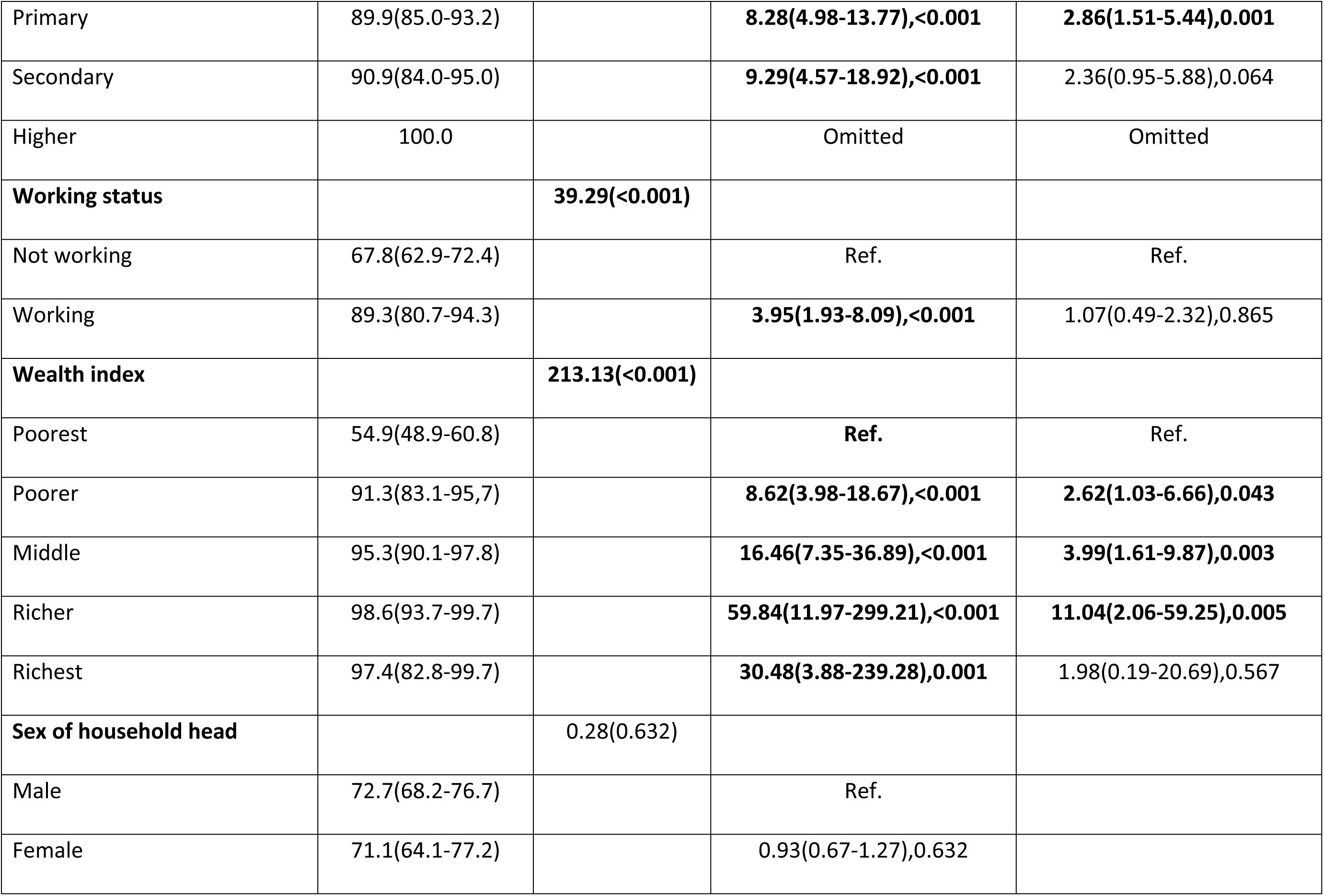
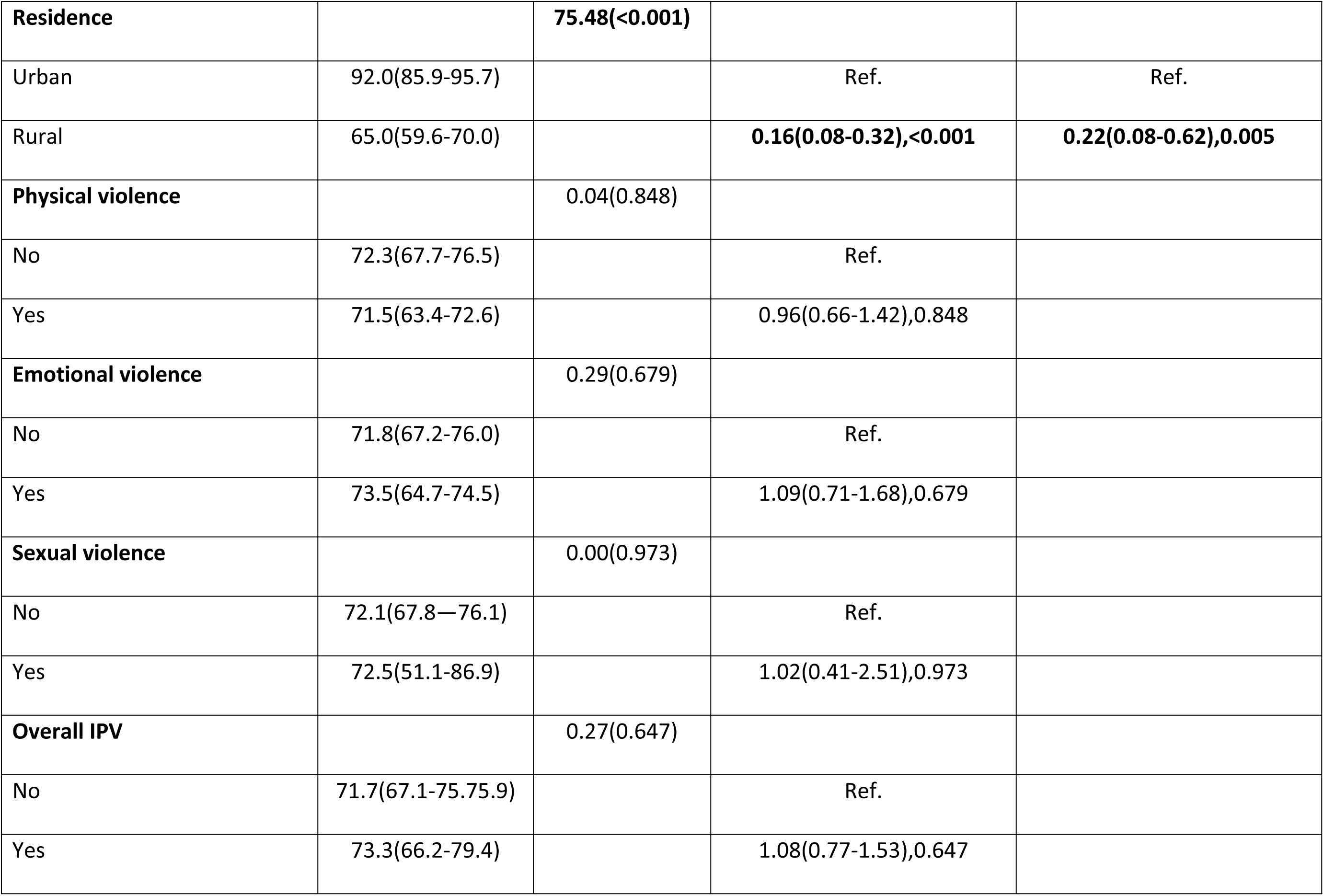
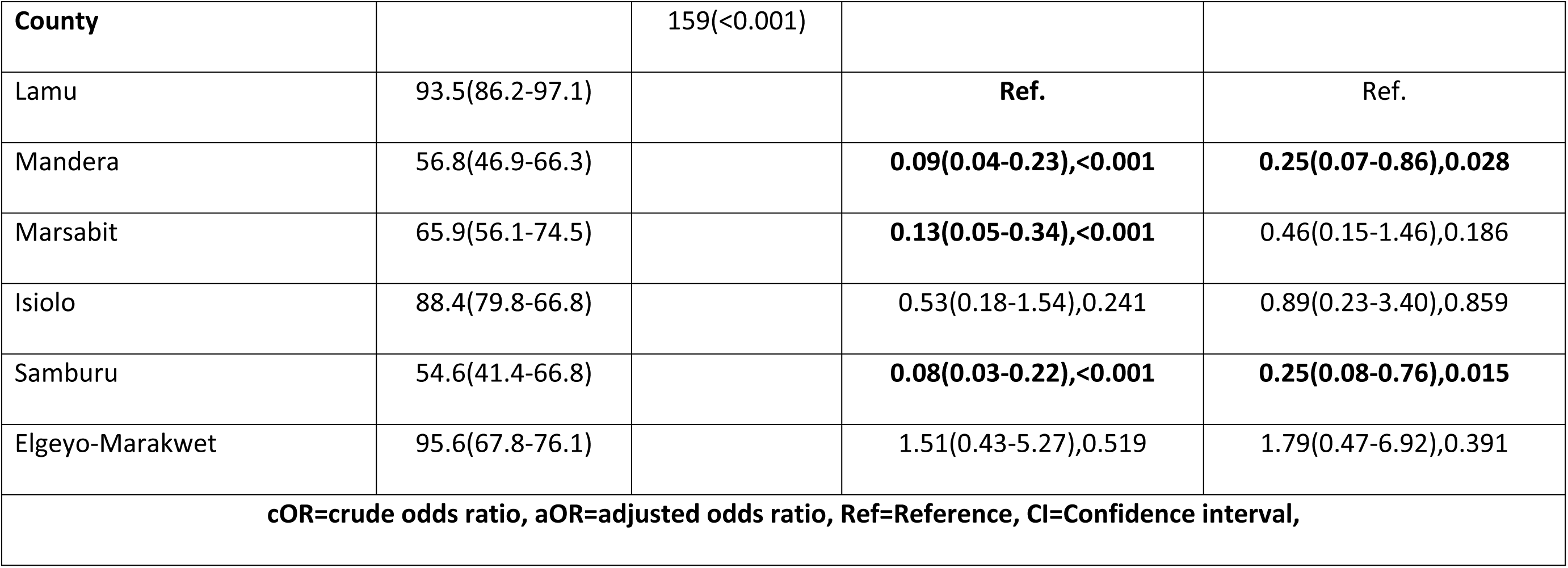
Bivariable and multivariable analysis of factors associated with the odds of skilled birth attendance.

| Variables | p(95% CI)<br>SBA | $\chi^2$ (p-value) | cOR (95% CI), p-value | aOR (95% CI), p-value |
| --- | --- | --- | --- | --- |
| **Age** |  | **18.07(0.037)** |  |  |
| 15-19 | 66.9(53.9-77.7) |  | Ref. |  |
| 20-24 | 79.3(72.5-84.8) |  | 1.89(0.99-3.62),0.053 |  |
| 25-29 | 76.7(70.9-81.7) |  | 1.64(0.88-3.05),0.122 |  |
| 30-34 | 66.6(58.3-74.0) |  | 0.98(0.51-1.93),0.976 |  |
| 35-39 | 67.6(57.3-76.8) |  | 1.04(0.56-1.93),0.912 |  |
| 40-44 | 62.0(44.4-77.0) |  | 0.81(0.33-2.01),0.647 |  |
| 45-49 | 68.5(32.6-90.7) |  | 1.08(0.21-5.48),0.929 |  |
| **Educational level** |  | **212.14(<0.001)** |  |  |
| None | 51.7(45.1-58.2) |  | Ref. | Ref. |
| Primary | 89.9(85.0-93.2) |  | **8.28(4.98-13.77),<0.001** | **2.86(1.51-5.44),0.001** |
| Secondary | 90.9(84.0-95.0) |  | **9.29(4.57-18.92),<0.001** | 2.36(0.95-5.88),0.064 |
| Higher | 100.0 |  | Omitted | Omitted |
| **Working status** |  | **39.29(<0.001)** |  |  |
| Not working | 67.8(62.9-72.4) |  | Ref. | Ref. |
| Working | 89.3(80.7-94.3) |  | **3.95(1.93-8.09),<0.001** | 1.07(0.49-2.32),0.865 |
| **Wealth index** |  | **213.13(<0.001)** |  |  |
| Poorest | 54.9(48.9-60.8) |  | Ref. | Ref. |
| Poorer | 91.3(83.1-95.7) |  | **8.62(3.98-18.67),<0.001** | **2.62(1.03-6.66),0.043** |
| Middle | 95.3(90.1-97.8) |  | **16.46(7.35-36.89),<0.001** | **3.99(1.61-9.87),0.003** |
| Richer | 98.6(93.7-99.7) |  | **59.84(11.97-299.21),<0.001** | **11.04(2.06-59.25),0.005** |
| Richest | 97.4(82.8-99.7) |  | **30.48(3.88-239.28),0.001** | 1.98(0.19-20.69),0.567 |
| **Sex of household head** |  | 0.28(0.632) |  |  |
| Male | 72.7(68.2-76.7) |  | Ref. |  |
| Female | 71.1(64.1-77.2) |  | 0.93(0.67-1.27),0.632 |  |
| **Residence** |  | **75.48(<0.001)** |  |  |
| Urban | 92.0(85.9-95.7) |  | Ref. | Ref. |
| Rural | 65.0(59.6-70.0) |  | **0.16(0.08-0.32),<0.001** | **0.22(0.08-0.62),0.005** |
| **Physical violence** |  | 0.04(0.848) |  |  |
| No | 72.3(67.7-76.5) |  | Ref. |  |
| Yes | 71.5(63.4-72.6) |  | 0.96(0.66-1.42),0.848 |  |
| **Emotional violence** |  | 0.29(0.679) |  |  |
| No | 71.8(67.2-76.0) |  | Ref. |  |
| Yes | 73.5(64.7-74.5) |  | 1.09(0.71-1.68),0.679 |  |
| **Sexual violence** |  | 0.00(0.973) |  |  |
| No | 72.1(67.8—76.1) |  | Ref. |  |
| Yes | 72.5(51.1-86.9) |  | 1.02(0.41-2.51),0.973 |  |
| **Overall IPV** |  | 0.27(0.647) |  |  |
| No | 71.7(67.1-75.75.9) |  | Ref. |  |
| Yes | 73.3(66.2-79.4) |  | 1.08(0.77-1.53),0.647 |  |
| **County** |  | 159(<0.001) |  |  |
| Lamu | 93.5(86.2-97.1) |  | **Ref.** | Ref. |
| Mandera | 56.8(46.9-66.3) |  | **0.09(0.04-0.23),<0.001** | **0.25(0.07-0.86),0.028** |
| Marsabit | 65.9(56.1-74.5) |  | **0.13(0.05-0.34),<0.001** | 0.46(0.15-1.46),0.186 |
| Isiolo | 88.4(79.8-66.8) |  | 0.53(0.18-1.54),0.241 | 0.89(0.23-3.40),0.859 |
| Samburu | 54.6(41.4-66.8) |  | **0.08(0.03-0.22),<0.001** | **0.25(0.08-0.76),0.015** |
| Elgeyo-Marakwet | 95.6(67.8-76.1) |  | 1.51(0.43-5.27),0.519 | 1.79(0.47-6.92),0.391 |
| **cOR=crude odds ratio, aOR=adjusted odds ratio, Ref=Reference, CI=Confidence interval,** |  |  |  |  |

Women in the "Richer" wealth quintile were 11 times more likely to deliver with a skilled attendant (aOR=11.04, 95% CI: 2.06–59.25, p=0.005) compared to the poorest women. Also, education remained the strongest determinant, significantly increasing the likelihood of SBA (aOR 2.86; 95% CI 1.51–5.44). Rural residence substantially reduced SBA utilisation (aOR 0.22; 95% CI 0.08–0.62). These geographic disparities across counties persisted after adjustment.

Intimate partner violence was not independently associated with skilled birth attendance or other maternal health indicators after controlling for sociodemographic factors, indicating that structural inequalities exerted the dominant influence on maternal service utilisation within conflict-affected counties.

## 4. Discussion

This study examined how sociodemographic factors structure maternal health service utilisation within Kenya’s conflict-affected counties. The findings demonstrate that maternal healthcare utilisation in these settings follows a rigid social gradient. Age, education, wealth, geographic residence, and county-level location were the dominant predictors of adequate antenatal care, facility delivery, and skilled birth attendance. In these fragile contexts, access to life-saving maternal care is therefore not merely a function of service availability, but of a woman’s capacity to mobilise socioeconomic resources in the face of insecurity and structural instability.

### 4.1 Age patterns and teenage pregnancies in conflict contexts

In our analysis, teenagers (15–19 years) were the least likely age group to achieve adequate antenatal care (ANC 4+).

The observed pattern is consistent with other evidence that teenagers and young mothers often experience lower ANC uptake and poorer continuity of maternal care, driven by stigma, low autonomy, poor knowledge of services, financial dependence, and limited mobility (18,19).

A recent systematic review on adolescent maternal health service utilisation showed evidence that adolescents’ use of maternal services is constrained by multiple barriers and that these barriers are likely intensified where systems are disrupted (20). Other reports from fragile and conflict-affected situations also show that younger women frequently face distinct and amplified constraints to sustaining ANC contacts, even when initial care is accessed (21).

Among Syrian refugees in Lebanon, Benage and colleagues documented major barriers in antenatal access in a displaced population, illustrating how conflict can reshape maternal care pathways for younger women (22). Peer-reviewed humanitarian literature also highlights that adolescent girls in crisis settings experience increased vulnerability to violence and exploitation. These are factors that can increase pregnancy risk and simultaneously reduce safe access to services (5,12). In conflict and displacement settings, adolescent risk is further compounded because insecurity disrupts protective social structures (like schooling and routine SRH services) while simultaneously increasing exposure to exploitation, coercion, and early unions in some contexts (9).

These findings posit treating teenage pregnancy and care as an equity priority in conflict-affected counties. Practical responses should explicitly lower barriers for teenagers by strengthening adolescent-friendly ANC care, stigma-free entry points, community follow-up mechanisms and safe referral/transport support in insecure rural areas; so that adolescent mothers are not systematically left behind during the very period when their risk is highest.

### 4.2 Education as a structural shield

Education emerged as the most consistent determinant across maternal health indicators. Educated women were significantly more likely to achieve adequate antenatal care, deliver in health facilities and access skilled birth attendance.

This finding aligns with Alie and Manyeh (23,24) who observed that maternal education provides the agency and knowledge required to navigate difficult health environments. Other authors posit that in conflict zones where security conditions fluctuate, educated women may possess a higher capacity for risk-assessment and information processing, enabling them to identify safe means of reaching clinics (25). Authors working in IDP camps in Sudan also reported a strong association between women’s level of education and maternal health seeking behaviours and choices. (26)

In insecure environments where transport routes fluctuate and information channels are unstable, the ability to assess risk and mobilise resources becomes critical. Studies from fragile and conflict-affected contexts similarly demonstrate that maternal education strengthens agency (8,10). It appears that individual capabilities probably become more decisive as systems destabilise.

The magnitude of effect observed in this study suggests that education does not merely increase utilisation incrementally; rather, it modifies a woman’s survival prospects in structurally constrained settings (10). These findings reinforce the centrality of female secondary education as a maternal survival strategy in fragile regions.

### 4.3 The wealth gradient during conflict

Another striking inequity observed was the wealth gradient in facility delivery and skilled birth attendance. Women in higher wealth quintiles were several times more likely to deliver in health facilities and with skilled attendants compared to those in the poorest quintile.

This suggests that in Kenya’s conflict counties, safe motherhood is becoming a costly economic commodity. Wealth appears to be protective with regards to women’s ability to seek care from skilled attendants (25).

In stable settings, free maternity policies (such as Linda Mama program) had been created to remove financial barriers in Kenya. However, as noted by Ziegler and collaborators, in conflict-affected regions in DRC, the hidden costs of care for example safe transportation and security; escalate dramatically leading to less than expected outputs in care seeking behaviours (27). The data suggests that women’s survival depends intricately on their ability to pay for different commodities. Only women with significant household wealth can afford the payments required to navigate insecure corridors to reach a facility (10). In conflict-affected counties, this gradient likely reflects more than routine user fees. Insecurity generates hidden and indirect costs: safer transportation, security risks, opportunity costs of travel, and potential informal payments (28,29).

### 4.4 The effect of rurality

Rural dwellers were 78% less likely to utilize skilled birth attendance (aOR=0.22) and 68% less likely to deliver in a health facility (aOR=0.32) compared to urban women in the same counties. In conflict settings, rurality is not just a measure of distance but also a measure of isolation from security and state protection (30). Other authors in sub Saharan Africa also found that living in a rural area significantly affected ANC attendance and care seeking behaviours (31,32)

In addition, according to the "Three Delays" framework by Thaddeus and Maine in 1994, conflict would act as a distance multiplier. Insecurity disrupts rural transport networks and leads to the retraction of skilled health workers to safer urban hubs (33,34). Consequently, for a rural woman in Samburu or Elgeyo Marakwet, the second delay (reaching care) becomes insurmountable during periods of active conflict, resulting in a reliance on traditional birth attendants despite the known risks in these regions.

### 4.5 County-level differences

Significant inter-county differences persisted across maternal health indicators even after adjustment for individual sociodemographic factors. This heterogeneity highlights an important point: conflict-affected counties are not homogeneous environments (35). Other Kenyan authors also reported disparities in maternal health indicators based on sub national differences (36)

Variation may reflect differences in intensity of insecurity, infrastructure stability, humanitarian presence, health system investment, and local governance. The persistence of county effects indicates that structural and contextual factors operate beyond individual-level characteristics(37). This reinforces the need for sub-national planning. National averages conceal meaningful variation, and policy responses must therefore be tailored to county-specific realities rather than applied uniformly across fragile regions.

### 4.6 The role of Intimate Partner Violence (IPV)

Notably, no form of Intimate Partner Violence (IPV) was significantly associated with maternal care indicators in our adjusted models. This finding contrasts with broader studies such as (38) but aligns partially with (39) who found no significant link between IPV and ANC in other low and middle income countries.

In Kenya’s conflict counties, this absent role of IPV may be explained by two factors. First, the underlying effect of extreme poverty, the barriers imposed by war and geographic isolation, poverty may be so overwhelming that they dwarf the effect of interpersonal violence on service utilization. Second, as suggested by (40) women experiencing violence may actually attend ANC *more* frequently as a safety-seeking behaviour or with the primary intention of managing injuries resulting from abuse. The lack of significance should not be interpreted as a lack of burden, but rather as evidence that structural determinants (age/wealth/education) are the primary influencers of survival in fragile states.

### 4.6 Strengths and Limitations

The primary strength of this study is the linkage of the 2022 KDHS with ACLED conflict metrics, providing an objective conflict classification. However, several limitations must be noted. As a secondary analysis of survey data, the study cannot establish temporal causality. Furthermore, DHS data on IPV is subject to underreporting due to the intense cultural stigmatization and learned helplessness often prevalent in patriarchal communities. Finally, while ACLED provides a fatality-based classification, it may not capture the fear of violence which can suppress service utilization even in the absence of active fatalities.

## 5. Conclusion

In Kenya’s conflict-affected counties, maternal health indicators are structured by a pronounced sociodemographic gradient, with teenagers, rural women, the poorest, and the least educated facing the greatest disadvantage. Teenagers are unlikely to attain adequate ANC. Education and wealth are protective while rurality and geography amplify exclusion. Equity-focused strategies should prioritise teenage ANC continuity, removal of rural access barriers and sustained investment in girls’ education and women’s economic empowerment to improve maternal health indicators in fragile settings.

## 6. Recommendations

The study’s findings carry implications for achieving Sustainable Development Goal 3 (maternal health) and SDG 5 (gender equality) in conflict-affected regions. Policy responses should therefore prioritise:

- Explicitly lowering barriers for teenagers by strengthening adolescent-friendly ANC services, community referral mechanisms, and safe referral/transport support in insecure rural areas.
- Reduction of geographic and socioeconomic disparities.
- Expansion of female secondary education and economic empowerment.
- Community-based strategies to improve skilled birth availability and attendance.

## Data Availability

No data was generated by this study. The following existing data sources were used: Kenya DHS from MEASURE DHS available via https://dhsprogram.com/data/

## Acknowledgement

Thanks go to Amu Hubert for support during the data analysis

